# Metabolomics data improve 10-year cardiovascular risk prediction with the SCORE2 algorithm for the general population without cardiovascular disease or diabetes

**DOI:** 10.1101/2024.04.29.24306593

**Authors:** Ruijie Xie, Sha Sha, Lei Peng, Bernd Holleczek, Hermann Brenner, Ben Schöttker

**Affiliations:** Division of Clinical Epidemiology and Aging Research, German Cancer Research Center, Im Neuenheimer Feld 581, 69120 Heidelberg, Germany; Faculty of Medicine, Heidelberg University, 69115 Heidelberg, Germany; Saarland Cancer Registry, Neugeländstraße 9, 66117 Saarbrücken, Germany; Network Aging Research, Heidelberg University, Bergheimer Straße 20, 69115 Heidelberg, Germany

**Author notes:** **Laboratory or institution of origin:** Division of Clinical Epidemiology and Aging Research, German Cancer Research Center, 69120, Heidelberg, Germany. **Correspondence to:** Ben Schöttker, PhD, Im Neuenheimer Feld 581, 69120, Heidelberg, Germany.

## Abstract

**BACKGROUND:** The value of metabolomic biomarkers for cardiovascular risk prediction is unclear. This study aimed to evaluate the potential of improved prediction of the 10-year risk of major adverse cardiovascular events (MACE) in large population-based cohorts by adding metabolomic biomarkers to the novel SCORE2 model, which was introduced in 2021 for the European population without previous cardiovascular disease or diabetes.

**METHODS:** Data from 187,039 and 5,578 participants from the UK Biobank (UKB) and the German ESTHER cohort, respectively, were used for model derivation, internal and external validation. A total of 249 metabolites were measured with nuclear magnetic resonance (NMR) spectroscopy. LASSO regression with bootstrapping was used to identify metabolites in sex-specific analyses and the predictive performance of metabolites added to the SCORE2 model was primarily evaluated with Harrell’s C-index.

**RESULTS:** Thirteen metabolomic biomarkers were selected by LASSO regression for enhanced MACE risk prediction (three for both sexes, six male- and four female-specific metabolites) in the UKB derivation set. In internal validation with the UKB, adding the selected metabolites to the SCORE2 model increased the C-index statistically significantly (*P*<0.001) from 0.691 to 0.710. In external validation with ESTHER, the C-index increase was similar (from 0.673 to 0.688, *P*=0.042). The inflammation biomarker, glycoprotein acetyls, contributed the most to the increased C-index in both men and women.

**CONCLUSIONS:** The integration of metabolomic biomarkers into the SCORE2 model markedly improves the prediction of 10-year cardiovascular risk. With recent advancements in reducing costs and standardizing processes, NMR metabolomics holds considerable promise for implementation in clinical practice.

**Clinical Perspective:** *What Is New?:*

- Model derivation and internal validation was performed in the UK Biobank and external validation in the German ESTHER cohort. The novel nuclear magnetic resonance (NMR) spectroscopy derived metabolomics data set of the UK Biobank is 23 times larger than the previously largest study that aimed to improve a cardiovascular risk score by metabolomics.
- The large sample size allowed us, for the first time, to select metabolites specific for men and women. We selected 13 out of 249 metabolomic biomarkers and derived a new sex-specific algorithm on top of the SCORE2 model. Our results show that the predictive accuracy of the model extended by metabolomic biomarkers is significantly higher than the SCORE2 model.

*What Are the Clinical Implications?:*

- Our findings imply that metabolomics data improve the performance of the SCORE2 algorithms for a more accurate 10-year cardiovascular risk prediction in apparently healthy individuals.
- As metabolomic analyses became standardized and affordable by the NMR technology in recent years, these measurements have a translation potential for clinical routine.

## Introduction

Cardiovascular disease (CVD) remains a major global health challenge, being a leading contributor to both morbidity and mortality worldwide.^1^ Despite a period of decline in several countries, the number of CVD-related deaths is anticipated to increase, attributed to demographic changes and increasing prevalence of unhealthy lifestyles in younger generations.^2, 3^

Introduced in 2021, SCORE2 is a new sex-specific competing risk algorithm derived from a comprehensive analysis of nearly 700,000 individuals from prospective European cohorts to evaluate the 10-year risk of major adverse cardiovascular events (MACE) in individuals without CVD or diabetes.^4^ Despite the foundational clinical risk assessment provided by algorithms such as SCORE2, there remains a critical need to enhance the precision of predictions to ascertain an individual’s susceptibility to future cardiovascular events.^5, 6^

With advancements in assay techniques, metabolomics has rapidly gained prominence for its potential to uncover the complex interplay of genetic, environmental, and pathological factors in the progression of CVD.^7-9^ While some studies have investigated the prognostic value of metabolomics in cardiovascular risk prediction, the majority are limited by small sample size or a lack of external cohort validation.^10-12^ Additionally, whereas previous research often considered sex as a covariate, emerging evidence underscores the importance of selecting metabolomic biomarkers in sex-specific analyses with adequate sample sizes as a crucial strategy to enhance the model’s predictive capabilities.^13, 14^

This study aims to utilize data from two large European cohorts to select metabolomic biomarkers through a sex-specific approach and to assess whether these metabolomic biomarkers improve the predictive performance of the SCORE2 model for 10-year MACE risk assessment.

## Methods

### Study population

The UK Biobank (UKB) is a large prospective cohort with 502,493 participants aged 37 to 73 years recruited from 13 March 2006 to 1 October 2010 across 22 assessment sites in England, Scotland, and Wales.^15^

The ESTHER study is an ongoing population-based cohort study conducted in Saarland, Germany, with recruited individuals aged 50 to 75.^16, 17^ This recruitment occurred during standard health checks by general practitioners (GPs) from 1 July 2000 to 30 June 2002, resulting in a participant base of 9,940 individuals. Follow-ups were conducted every two to three years thereafter.

### In- and exclusion criteria

In the UKB and ESTHER cohort n=247,353 and n=8,308 study participants were randomly selected for metabolomics measurements, respectively. To ensure comparability in age distribution between two cohorts, participants not in the age range of 50-69 years were excluded (see **Supplemental Figure S1** for study flow-charts). We further excluded individuals with missing data on diabetes or those diagnosed with diabetes at the baseline. Additionally, we excluded participants with a prior history of myocardial infarction or stroke before baseline or those lacking precise dates for these cardiovascular events. Finally, we included n=187,039 and n=5,578 individuals with measured metabolomics data from the UKB and ESTHER study, respectively.

### Measurement of metabolomic biomarkers

Nightingale Health utilized a high-throughput nuclear magnetic resonance (NMR) metabolomics platform to analyze plasma samples in UKB participants and serum samples from the ESTHER cohort. This analytical process facilitated the identification of 250 metabolomic biomarkers. These biomarkers encompassing a range of molecules such as lipids, fatty acids, amino acids, ketone bodies, and other critical low-molecular-weight compounds.^18^ Nevertheless, glycerol was omitted from the analysis because it could not be measured in most samples in both cohorts, reducing the number of analyzed metabolites to 249. Details regarding these metabolites, including their names and completeness, are presented in **Supplemental Table S1**.

### Variables of the SCORE2 model

The SCORE2 model, designed for adults without diabetes aged 40 to 69, includes age, sex, high-density lipoprotein cholesterol (HDL-C), total cholesterol, systolic blood pressure (SBP), and smoking status.^4^

For both cohorts, standardized questionnaires were utilized to collect data on age, sex, and smoking status (current or not). HDL-C and total cholesterol were measured with an enzymatic method in both cohorts (UKB: Beckman Coulter AU5800; ESTHER: Cobas 8000 C701). SBP measurements were conducted by automated reading of the Omron device at the left upper arm in the UKB. In the ESTHER study, SBP measurement methods varied by the GPs and the results were documented in the physician’s medical conditions report of the health check-up.^19^

### Outcome ascertainment

The primary endpoint was MACE, defined as a composite of cardiovascular death, non-fatal myocardial infarctions, and non-fatal strokes, with a detailed definition provided in **Supplemental Table S2**. This was aligned with the criteria of the SCORE2 model.^4^ Except for a broader classification of non-fatal strokes in the ESTHER study due to the inability to differentiate stroke subtypes in this cohort.

As outlined in previous studies,^20^ the ESTHER study participants recorded incidents of myocardial infarction and stroke through standardized surveys conducted 2, 5, 8, and 11 years after baseline. These self-reported incidents were confirmed via questionnaires sent to the participants’ GPs, with 89% of myocardial infarction and 91% of stroke cases used in this analysis verified by the GPs. Additionally, a vital status check was conducted at registration offices, and death certificates from local health authorities were obtained for deceased participants.

In the UKB, occurrences of non-fatal myocardial infarctions and strokes were ascertained from primary care records or hospital episode statistics. Dates and causes of death were established using death registries from the National Health Service Information Centre in England and Wales, and the National Health Service Central Register in Scotland.

Study participants were censored at the first occurrence of a non-fatal myocardial infarction, stroke, or death in the first ten years after baseline or when the maximum follow-up time of ten years was reached.^4^

### Statistical analyses

#### General remarks

All analyses were conducted using R software, version 4.3.0 (R Foundation for Statistical Computing, Vienna, Austria), with statistical significance set at *P*-values<0.05 for two-sided tests. Missing values were imputed using the random forest imputation method, as implemented in the r-package *missForest* (version 1.5).^21^ Most variables of the SCORE2 model and the NMR metabolites were complete, and the proportion of imputed missing values varied from 0 to 11.1% (**Supplemental Table S1**).

### Metabolites selection and model derivation

Initially, all metabolite concentrations underwent log-transformation to approximate normal distributions for analysis. These measures were then scaled to standard deviation (SD) units independently within each cohort. The UKB dataset was partitioned into a derivation set (70%) and an internal validation set (30%). The ESTHER study served as the external validation cohort. The derivation set was used for metabolite selection and model derivation.

The Least Absolute Shrinkage and Selection Operator (LASSO), a regularization technique adept at identifying strong predictors among high-dimensional and correlated predictors, was implemented via the r-package *glmnet* (version 4.1-7).^22^ We employed the ten-fold cross-validation to determine the optimal tuning parameter λ for LASSO, based on the lowest model validation error. A bootstrap LASSO approach was then undertaken, involving the creation of 1000 derived sets with the LASSO procedure applied to each resampled dataset.^23^ Metabolites selected by LASSO in at least 95% of instances were designated as our metabolites of interest. The threshold was adopted from a previous study to enhance model generalization and mitigate overfitting.^24^ The selected metabolites were subsequently incorporated into the SCORE2 model to construct new sex-specific competing risk algorithms using the r-package *cmprsk* (version 2.2-11).

### Predictive model performance validation

The predictive performance of the derived model was validated by using 30% of the UKB as the internal validation cohort and the ESTHER cohort as the external validation cohort. The incremental discrimination contributed by each metabolite was evaluated based on the internal validation results.

Discriminative ability was assessed using Harrell’s C-index, adjusted for competing risks (via the r-package *riskRegression*, version 2023.03.2).^25^ Furthermore, the net reclassification index (NRI) and the integrated discrimination improvement (IDI) were evaluated for the final model (using the r-package *nricens*, version 1.6).^26^ Pre-specified CVD risk categories (0–5%, >5– 10%, and >10%) were employed for the NRI to indicate the proportion of individuals correctly reclassified compared with the SCORE2 model. Model calibration was examined by plotting observed MACE event rates against predicted event rates across deciles of absolute predicted risk.

### Associations of selected metabolites with MACE

To report hazard ratios (HRs) and 95% confidence intervals (CIs) for the selected metabolites (per one SD increment) associated with 10-year MACE incidence in male and female study participants, metabolites were individually added to Cox proportional hazards regression models in both the internal validation cohort and the external validation cohort, respectively. These models were additionally adjusted for the SCORE2 model variables, using the r-package *survival* (version 3.5-5). The Benjamini-Hochberg procedure was applied to control the false discovery rate (FDR) in this analysis.

## Results

### Baseline characteristics and MACE case numbers

**Table 1** summarizes the characteristics of the 187,039 included participants from the UKB and the 5,578 included participants from the ESTHER study for all variables included in the SCORE2 model. The UKB participants had an average age of 59.8 years (SD 5.4) and 56.1% were male. Over a follow-up time limited to 10 years, 9,396 MACE events were recorded in the UKB. The ESTHER participants had a mean age of 60.0 years (SD 5.5), and 58.3% of the participants were male. In total, 474 MACE events were recorded in the first 10 years of follow-up in ESTHER.

**Table 1.** Baseline characteristics of selected participants from the UK Biobank and ESTHER study.

| Baseline characteristics | UK Biobank (n=187,539) | ESTHER (n= 5,578) |
| --- | --- | --- |
| Male sex, N (%) | 105,301 (56.1) | 3,253 (58.3) |
| Age (years), mean (SD) | 59.8 (5.4) | 60.0 (5.5) |
| Current smoker, N (%) | 17,307 (9.2) | 897 (16.1) |
| Systolic blood pressure (mmHg), mean (SD) | 142.4 (19.7) | 138.1 (19.0) |
| Total cholesterol (mmol/L), mean (SD) | 4.8 (0.9) | 5.8 (1.1) |
| HDL cholesterol (mmol/L), mean (SD) | 1.4 (0.3) | 1.4 (0.3) |

**Abbreviations:** HDL, high-density lipoprotein; SD, standard deviation.

### Associations of metabolomic biomarkers with MACE

A total of 13 metabolites were selected to enhance MACE risk prediction in the SCORE2 model in the derivation cohort by LASSO analyses and bootstrapping with a frequency of more than 950 times in the 1000 replicate selection (**Supplemental Table S3**). These metabolites include three that improved MACE risk prediction in both sexes — albumin, GlycA (glycoprotein acetyls), and IDL-CE-pct (the cholesteryl esters percentage of total lipids in intermediate-density lipoprotein). Additionally, four metabolites were specifically predictive in females - acetoacetate, LA-pct (the linoleic acid percentage of total fatty acids), L-LDL-TG-pct (the triglycerides percentage of total lipids in large low-density lipoprotein), and valine - and six were exclusive to males: acetate, L-VLDL-TG-pct (the triglycerides percentage of total lipids in large very-low-density lipoprotein), Omega-3-pct (the Omega-3 fatty acids percentage of total fatty acids), S-HDL-CE (cholesteryl esters in small high-density lipoprotein), VLDL-size (average diameter of very-low-density lipoprotein particles), and XL-HDL-FC (free cholesterol in very large high-density lipoprotein).

Figure 1 shows the associations of the 13 selected metabolites with MACE in multivariate Cox regression models adjusted for SCORE2 model variables assessed separately in males and females in the internal validation set of the UKB. GlycA, L-LDL-TG-pct and VLDL-size were positively, and LA-pct, Omega-3-pct, albumin, IDL-CE-pct, L-VLDL-TG-pct and S-HDL-CE were inversely, statistically significantly associated with MACE at an FDR-adjusted *P*<0.05 level in both sexes. Acetate was only statistically significant in males and XL-HDL-FC was only statistically significant in females with an FDR-adjusted p<0.05. Due to the lower statistical power of the ESTHER study, of the 13 selected metabolites, only GlycA, acetate, IDL-CE-pct and L-LDL-TG-pct were statistically significantly associated with MACE in women (**Supplemental Figure S2**).

**Figure 1.**
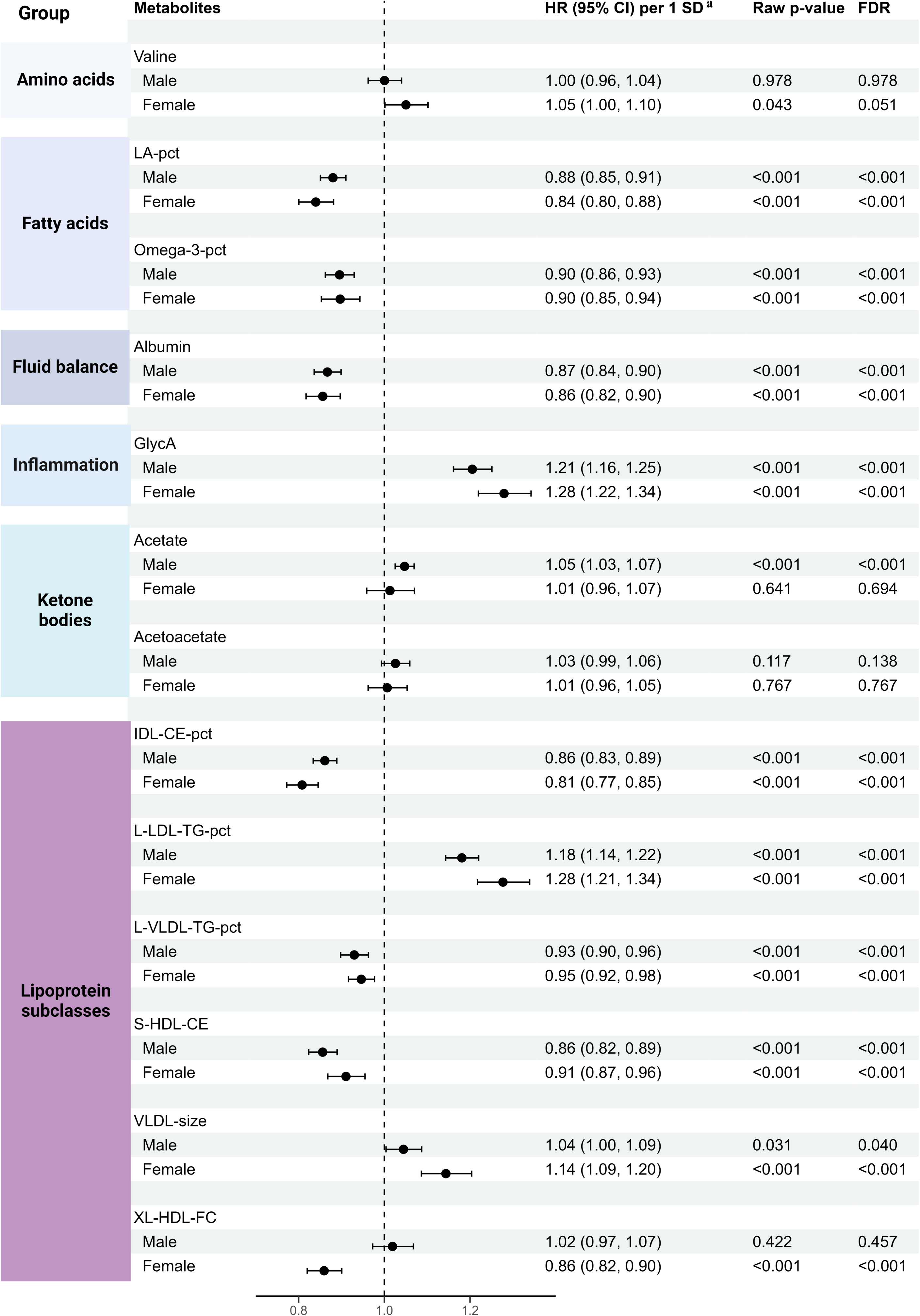
Associations between selected metabolites and major cardiovascular events across sexes in the internal validation cohort (30% of UK Biobank, N=56,112) **Abbreviations:** CI, confidence interval; FDR, false discovery rate; GlycA, glycoprotein acetyls; IDL-CE-pct, the cholesteryl esters percentage of total lipids in intermediate-density lipoprotein; LA-pct, the linoleic acid percentage of total fatty acids; L-LDL-TG-pct, triglycerides to total lipids in large low-density lipoprotein percentage; L-VLDL-TG-pct, the triglycerides percentage of total lipids in large very-low-density lipoprotein; Omega-3-pct, the Omega-3 fatty acids percentage of total fatty acids; SD, standard deviation; S-HDL-CE, cholesteryl esters in small high-density lipoprotein; VLDL-size, average diameter for very-low-density lipoprotein particles; XL-HDL-FC, free cholesterol in very large high-density lipoprotein. ^a^ The standard deviations of the metabolites acetate, albumin, GlycA, IDL-CE-pct, L-VLDL-TG-pct, Omega-3-pct, S-HDL-CE, VLDL-size, XL-HDL-FC, acetoacetate, LA-pct, L-LDL-TG-pct, and valine were 0.03 mmol/L, 3.35 mmol/L, 0.12 mmol/L, 2.31%, 5.84%, 1.58%, 0.05 mmol/L, 1.21 mmol/L,0.01 mmol/L, 0.01 mmol/L, 3.33mmol/L, 1.68%, and 0.04 mmol/L, respectively.

### MACE risk prediction by metabolomic biomarkers

**Table 2** presents the predictive performance metrics of the SCORE2 model for 10-year MACE risk prediction, augmented by 13 metabolomic biomarkers. The coefficients for all variables in the extended risk model are detailed in **Supplemental Table S3**. In the derivation set, the integration of these metabolites statistically significantly bolstered the C-index of the SCORE2 model (*P*<0.001) from 0.691 (95% CI: 0.691, 0.692) to 0.712 (95% CI: 0.712, 0.712) for the total sample of women and men. This improvement was more pronounced in males (+0.034) than in females (+0.021), with statistical significance observed in both sexes (*P*<0.001). Internal validation within the UKB cohort confirmed these findings, showcasing very similar C-index values. Moreover, the enhanced model demonstrated significant gains in reclassification statistics for both sexes, evidenced by statistically significant improvements in NRIs and IDIs (*P*<0.01). Calibration of both the SCORE2 and extended models exhibited satisfactory performance in the internal validation set, as depicted in **Supplemental Figure S3**.

**Table 2.** Metrics of the predictive performance of the SCORE2 model for 10-year MACE risk without and with extension by metabolites.

| Metrics | Male * | Female † | Overall |
| --- | --- | --- | --- |
| **Derivation set (70% of UK Biobank)** |  |  |  |
| **Total sample size (N=130,927)/MACE case number (N=6,573)** |  |  |  |
| C-Statistics (SCORE2) | 0.632 (0.630, 0.635) | 0.688 (0.686, 0.690) | 0.691 (0.691, 0.692) |
| C-Statistics (+Metabolites) | 0.666 (0.664, 0.668) | 0.709 (0.707, 0.711) | 0.712 (0.712, 0.712) |
| *P*-values for C-Statistics comparisons | **<0.001** | **<0.001** | **<0.001** |
| **Internal validation set (30% of UK Biobank)** |  |  |  |
| **Total sample size (N=56,112)/MACE case number (N=2,823)** |  |  |  |
| C-Statistics (SCORE2) | 0.638 (0.625, 0.650) | 0.686 (0.670, 0.702) | 0.691 (0.688, 0.694) |
| C-Statistics (+Metabolites) | 0.665 (0.652, 0.677) | 0.709 (0.693, 0.724) | 0.710 (0.707, 0.713) |
| *P*-values for C-Statistics comparisons | **<0.001** | **<0.001** | **<0.001** |
| NRI continuous (%) | **13.1 (6.1, 16.9)** | **12.9 (1.7, 21.9)** | **10.5 (1.5, 15.1)** |
| NRI categorical total (%) | **8.4 (4.8, 12.0)** | **3.9 (1.0, 8.8)** | **6.7 (4.8, 9.1)** |
| NRI categorical events (%) | **3.5 (1.2, 6.3)** | 3.4 (-1.1, 8.6) | **5.1 (3.1, 7.7)** |
| NRI categorical non-events (%) | **4.9 (1.8, 6.9)** | 0.5 (-1.1, 2.5) | **1.6 (1.1, 2.1)** |
| IDI | **0.009 (0.005, 0.012)** | **0.007 (0.003, 0.011)** | **0.007 (0.003, 0.009)** |
| **External validation set (ESTHER)** |  |  |  |
| **Total sample size (N=5,578)/MACE case number (N=474)** |  |  |  |
| C-Statistics (SCORE2) | 0.602 (0.562, 0.642) | 0.715 (0.680, 0.749) | 0.673 (0.647, 0.698) |
| C-Statistics (+Metabolites) | 0.629 (0.589, 0.669) | 0.724 (0.690, 0.757) | 0.688 (0.663, 0.713) |
| *P*-values for C-Statistics comparisons | 0.099 | 0.317 | **0.042** |
| NRI continuous (%) | **13.0 (0.6, 20.7)** | 7.8 (-13.7, 23.3) | **16.2 (1.9, 25.8)** |
| NRI categorical total (%) | 3.4 (-4.0, 11.9) | 7.7 (-1.0, 14.0) | 8.0 (-2.0, 14.0) |
| NRI categorical events (%) | -0.1 (-7.4, 6.9) | 4.8 (-3.1, 8.2) | 3.2 (-2.6, 6.7) |
| NRI categorical non-events (%) | 3.5 (-5.4, 12.7) | 2.9 (-2.7, 8.1) | 4.8 (-4.2, 11.6) |
| IDI | **0.010 (0.000, 0.018)** | 0.006 (-0.008, 0.023) | **0.011 (0.002, 0.023)** |

**Abbreviations:** IDI, integrated discrimination improvement; NRI, net reclassification improvement.
\* Metabolites that were included in the combined model for male were acetate, albumin, GlycA, IDL-CE-pct, L-VLDL-TG-pct, Omega-3-pct, S-HDL-CE, VLDL-size, and XL-HDL-FC.
† Metabolites that were included in the combined model for female were acetoacetate, albumin, GlycA, IDL-CE-pct, LA-pct, L-LDL-TG-pct, and valine.

In the external validation using the ESTHER cohort, the model displayed also a pronounced improvement in discrimination (C-index increase from 0.673 to 0.688; **Table 2**), reaching statistical significance in the overall sample of men and women (*P*=0.042). This improvement was more pronounced in males (+0.027) than in females (+0.009) but was not statistically significant in either of the sexes (*P*>0.05). Regarding reclassification, although the categorical NRI did not show significant improvement (total categorical NRI: 8.0, 95% CI: -2.0, 14.0), both the continuous NRI and IDI exhibited significant enhancements in the total cohort (continuous NRI: 16.2, 95% CI: 1.9, 25.8; IDI: 0.011, 95% CI: 0.002, 0.023). These findings still illustrate the superior performance of the extended model over the SCORE2 model in risk reclassification. However, in the external validation set, both the SCORE2 and extended models demonstrated suboptimal calibration by underestimating the observed risk in both male and female participants (**Supplemental Figure S4**).

Figure 2 illustrates the incremental enhancement in the C-statistic achieved by sequentially incorporating the 13 selected metabolites into sex-specific models in the internal validation set. Notably, in males, the inclusion of acetate, GlycA, IDL-CE-pct, Omega-3-pct, S-HDL-CE, and XL-HDL-FC significantly improved the models’ discriminative capabilities. While albumin and L-VLDL-TG-pct also showed improvement in males, these changes were not statistically significant. In females, the addition of GlycA, IDL-CE-pct, and L-LDL-TG-pct significantly augmented discrimination. Additionally, albumin and LA-pct contributed to an appreciable, albeit non-significant, increase in the C-index. **Supplemental Figure S5** shows the results of the same analysis for the external validation. All 9 metabolites led to a non-significantly improved C-index in men, and in women, 4 out of 7 showed a non-significant increase in the C-index. Taking the results for men and women together, both in internal validation and external validation, the inflammation biomarker GlycA contributed the largest share in C-index increase of all selected metabolites.

**Figure 2.**
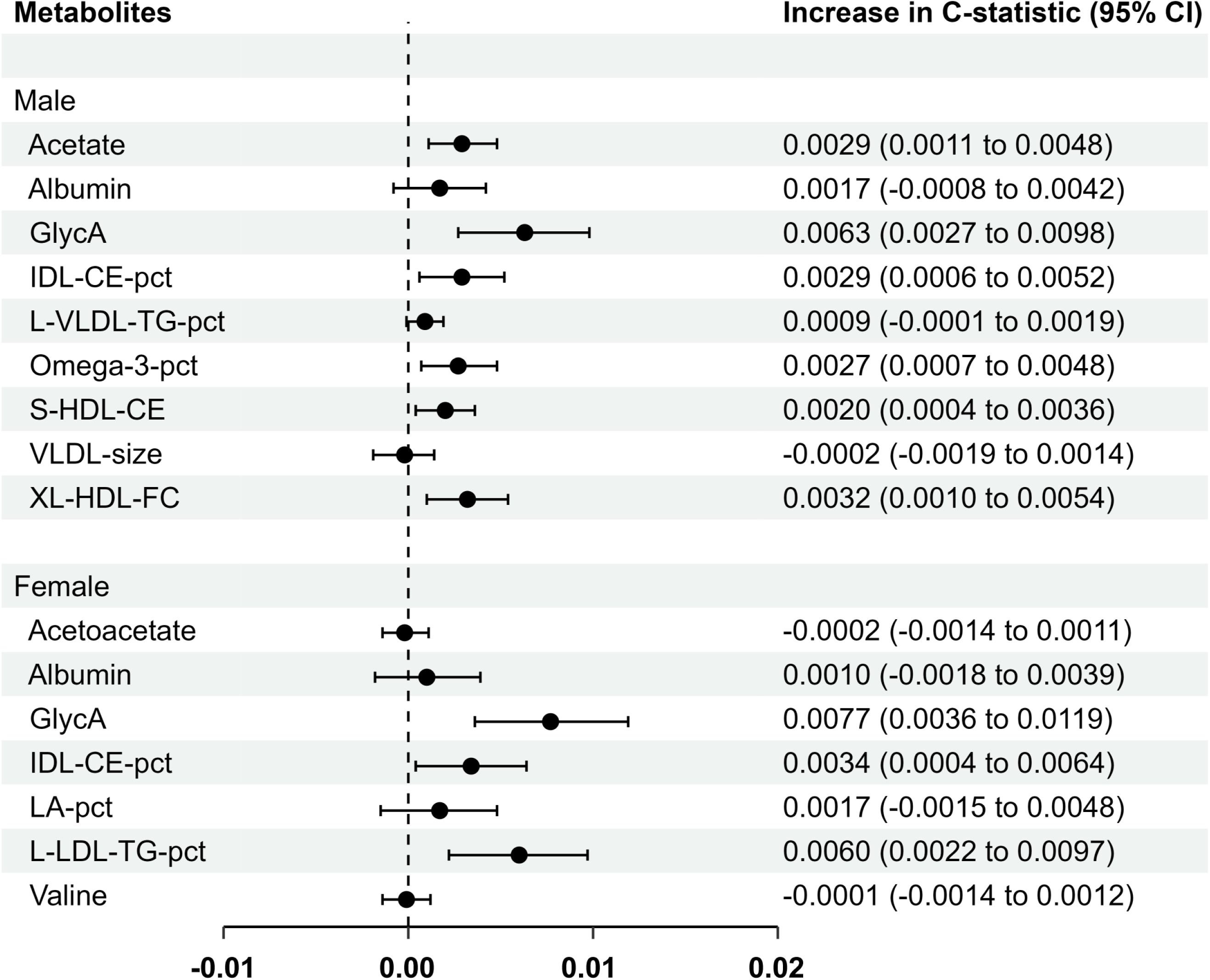
The incremental discrimination of each metabolite for the model after the selected metabolites were added to SCORE2 separately in different sexes in the internal validation cohort (30% of UK Biobank, N=56,112) **Abbreviations:** CI, confidence interval; GlycA, glycoprotein acetyls; IDL-CE-pct, the cholesteryl esters percentage of total lipids in intermediate-density lipoprotein; LA-pct, the linoleic acid percentage of total fatty acids; L-LDL-TG-pct, triglycerides to total lipids in large low-density lipoprotein percentage; L-VLDL-TG-pct, the triglycerides percentage of total lipids in large very-low-density lipoprotein; Omega-3-pct, the Omega-3 fatty acids percentage of total fatty acids; S-HDL-CE, cholesteryl esters in small high-density lipoprotein; VLDL-size, average diameter for very-low-density lipoprotein particles; XL-HDL-FC, free cholesterol in very large high-density lipoprotein.

## Discussion

### Summary of the findings

Using large-scale data from two European cohorts of middle-aged and older participants, we derived and validated an algorithm combining the SCORE2 model and 13 metabolites for improving 10-year cardiovascular risk prediction. In internal and external validation, the C-index of the SCORE2 model significantly increased by 0.019 and 0.015, respectively. In addition, the reclassification ability of the SCORE2 model was significantly improved in both internal and external validation. In terms of calibration, the performance of the SCORE2 and its extended model showed good calibration in the internal validation in the UKB, but the external validation in the German ESTHER study revealed an underestimation of risk in both the original and extended model. Thus, calibration of the models to the underlying CVD risk in specific regions is still needed.

### Comparison with previous studies & novelty

The high-dimensional nature of metabolomics data dictates that sample size is a crucial factor for accurately selecting the most predictive metabolites for health outcomes. Compared to previous studies, this study represents the largest sample size to date in evaluating cardiovascular risk models with metabolomic biomarkers. A recent systematic review and meta-analysis, which included and analyzed 21 studies using metabolomics to assess cardiovascular risk, encompassing a total of 54,337 participants, which is less than one third of the sample size used from the UKB alone in this work (n=187,039). The meta-analysis revealed that incorporating metabolites into the prediction model resulted in an average increase of 0.04 in the C-index,^12^ which was higher than in our results. This might be explained by more metabolites measured. A narrow focus on specific categories of metabolomic biomarkers may limit the accuracy of risk prediction.^27, 28^ However, previous studies were constrained by their relatively small sample sizes, with the largest cohort consisting of 8,101 participants,^29-32^ which raises concerns regarding to the robustness of their models and, coupled with a widespread absence of external model validation, restricts the generalizability of these models.^33, 34^

Recent evidence has highlighted that there are sex differences in both cardiovascular disease risk and metabolomic biomarker blood concentrations, suggesting that the derivation of sex-specific risk models is a valuable approach to enhance predictive accuracy.^13, 35^ Previous studies typically included sex as a covariate in the model derivation process to select metabolites, rather than selecting metabolites based on sex for males and females separately. The primary reason might be that sex-specific analyses are much more demanding in terms of sample size.

### Biological mechanisms of selected metabolites for MACE risk prediction

Three metabolites (albumin, GlycA and IDL-CE-pct) were chosen to enhance MACE risk prediction for both males and females, four metabolites were additionally selected for females (acetoacetate, LA-pct, L-LDL-TG-pct, and valine), and six metabolites were specifically chosen for males (acetate, L-VLDL-TG-pct, Omega-3-pct, S-HDL-CE, VLDL-size and XL-HDL-FC). Albumin serves as a biomarker of nutritional status and acts as a critical indicator of systemic inflammation and renal health, both of which are pivotal in the assessment and management of cardiovascular risk.^36^ GlycA reflects the glycosylation status of acute phase proteins and thus is a pro-inflammatory biomarker. Inflammatory processes are crucial in the development and progression of cardiovascular diseases.^37^ Acetoacetate is a type of ketone body and has been shown to enhance contractile performance and beta-adrenergic inotropism in stunned myocardium, as indicated by its circulating levels. Clinical research suggests that in non-diabetic populations, a significant association exists between the presence of acetoacetate and the occurrence of heart failure.^38^ Omega-3-pct and LA-pct may reduce cardiovascular risks by modulating lipid profiles and inflammatory markers.^39, 40^ Valine is a branched-chain amino acid (BCAA) and has been recognized for its ability to indicate cardiac metabolic risks independently of critical factors such as sex and BMI.^41^ Acetate is the most abundant endogenously produced short-chain fatty acid, is associated with weight loss, enhanced insulin sensitivity, and reduced cardiovascular risk.^42^ Lipoprotein components, such as IDL-CE-pct and L-VLDL-TG-pct, represent the intricate dynamics of lipid content and particle size within lipoprotein subclasses, which are crucial in the development and progression of atherosclerosis.^43-46^

### Strengths and limitations

Our investigation stands out for its considerable cohort size, comprising 187,039 participants in the UKB and 5,578 in the ESTHER study, rendering it the largest metabolomics analysis dedicated to cardiovascular risk prediction to date. Additionally, this study is pioneering in selecting metabolomic biomarkers for cardiovascular risk prediction separately by sex. Furthermore, model calibration was performed for a low (UK) and intermediate cardiovascular risk region (Germany). Although the results of our model calibration were not satisfactory in external validation, two recent studies using the SCORE2 model in other European cohorts (the EPIC-Norfolk study and a cohort based on a Netherland’s population) have also shown poor model calibration, suggesting that recalibration of model risk is warranted in external cohorts.^47, 48^

Our study also has limitations. The ESTHER study may not be the most ideal external validation cohort for our model due to differences with the UKB, particularly in the following aspects: the determination of non-fatal strokes cannot exclude hemorrhagic strokes; and regional differences in cardiovascular risk (low in UK and intermediate in Germany). Most blood samples in the UKB cohort were collected in a non-fasting state (96.6%) in plasma samples, whereas most samples in the ESTHER study were collected in a fasting state in serum samples (89.4%). However, a comparison of the levels of the 13 selected metabolites by fasting status in the UK Biobank and ESTHER study shows that the differences in metabolite concentrations between fasting and non-fasting individuals are minimal in both cohorts (see **Supplement Table S4**). This is in agreement with a previous study, which has shown that the duration of fasting has little impact on the variability of these biomarkers.^49^ However, despite these cohort differences, the overall consistency in risk prediction results between the two cohorts provides support for the robustness of the derived model. Although this study may be generalizable to low-to-intermediate risk regions in Europe, larger validation cohorts are needed, which should originate not only from low to intermediate risk regions in Europe but shall also validate the new risk model in populations from high and very high-risk regions.

### Potential clinical implications of the derived model

The inclusion of thirteen selected metabolomic biomarkers in the SCORE2 model has led to a significant improvement in the discrimination and reclassification of 10-year cardiovascular risk assessment. This modification not only bolsters the predictive capacity of the model but also maintains the simplicity and efficiency of traditional cardiovascular risk assessments, which typically involve only a brief physical examination, patient interview, and blood analysis.

Incorporating metabolomics into standard clinical evaluations undoubtedly raises expenses. However, it is crucial to recognize that NMR presents a more economical and uniform approach for metabolomic analysis compared to liquid chromatography-mass spectrometry/mass spectrometry (LC-MS/MS).^50^ The ability of commercial organizations like Nightingale Health, which can efficiently handle large volumes of samples highlights the feasibility and potential for widespread application in clinical settings.

Prior to recommending routine implementation in clinical practice, it is imperative to conduct model validation specific to particular countries. Currently, our algorithm is primarily validated for the UK and Germany, and extending this validation to other regions is crucial for broader applicability. The differences in cardiovascular risk profiles and healthcare systems across countries necessitate such validation to ensure the model’s predictive performance and relevance in diverse settings.

## Conclusion

To conclude, our study demonstrated that integrating 13 metabolomic biomarkers from the Nightingale Health’s panel enhanced the predictive accuracy of the SCORE2 model for sex-specific 10-year cardiovascular risk assessment. Recent advancements in NMR technology have standardized and reduced the cost of metabolomic analyses, offering promising prospects for their integration into clinical practice. The enhancement of the SCORE2 model with 13 metabolites leads to more accurate cardiovascular risk stratification, a crucial component for devising personalized preventive strategies.

## Data Availability

Data from ESTHER is available upon reasonable request that is compatible with participants informed consent. Data from the UK Biobank (https://www.ukbiobank.ac.uk/) is available to bona fide researchers on application.

## Non-standard Abbreviations and Acronyms

BCAA: branched-chain amino acid;
CI: confidence intervals;
CVD: cardiovascular disease;
eGFR: estimated glomerular filtration rate;
ESTHER: Epidemiologische Studie zu Chancen der Verhütung, Früherkennung und optimierten Therapie chronischer Erkrankungen in der älteren Bevölkerung;
FDR: false discovery rat;
GlycA: glycoprotein acetyls;
GP: general practitioner;
HDL-C: high-density lipoprotein cholesterol;
HR: hazard ratio;
IDI: integrated discrimination improvement;
IDL-CE-pct: the cholesteryl esters percentage of total lipids in intermediate-density lipoprotein;
LA-pct: the linoleic acid percentage of total fatty acids;
LASSO: least absolute shrinkage and selection operator;
LC-MS/MS: Liquid Chromatography-Mass Spectrometry/Mass Spectrometry;
L-LDL-TG-pct: the triglycerides percentage of total lipids in large low-density lipoprotein;
L-VLDL-TG-pct: the triglycerides percentage of total lipids in large very-low-density lipoprotein;
MACE: major adverse cardiovascular events;
NMR: nuclear magnetic resonance;
NRI: net reclassification index;
Omega-3-pct: the Omega-3 fatty acids percentage of total fatty acids percentage;
SBP: systolic blood pressure;
SD: standard deviation;
S-HDL-CE: cholesteryl esters in small high-density lipoprotein;
UKB: UK Biobank;
VLDL-size: the average diameter for very low-density lipoprotein particles;
XL-HDL-FC: free cholesterol in very large high-density lipoprotein.

## ARTICLE INFORMATION

### Acknowledgements

We would like to thank all participants of the ESTHER and UKB cohorts as well as the GPs of the ESTHER study and the staff of the UKB assessment centers for their contribution to the studies this research is based on. The UKB analyses were conducted under application no. 101633.

### Supplemental Material

Figures S1–S5

Tables S1–S4

